# The changing epidemiology of shigellosis in Australia, 2001-2019

**DOI:** 10.1101/2022.05.03.22274596

**Authors:** Aaliya Ibrahim, Kathryn Glass, Deborah A Williamson, Ben Polkinghorne, Danielle J Ingle, Rose Wright, Martyn D Kirk

## Abstract

Shigellosis is an increasing cause of gastroenteritis in Australia, including prolonged outbreaks in remote Aboriginal and Torres Strait Islander (hereafter “First Nations”) communities and among men who have sex with men (MSM) in major cities. To determine associations between *Shigella* species and demographic and geographic factors we used negative binomial regression to analyse national case notifications of shigellosis from 2001 to 2019.

*S. sonnei* and *S. flexneri* accounted for 42% and 29% of cases, respectively. Nationally, notification rates increased from 2001 to 2019 with yearly incidence rate ratios of 1.04 (95% CI 1.02-1.07) for *S. boydii*, 1.05 (95% CI 1.04-1.06) for *S. sonnei* and 1.04 (95% CI 1.04-1.06) for *S. flexneri*. Children aged 0-4 years had the highest burden of infection for *S. flexneri, S. sonnei* and *S. boydii*; and males had a higher notification rate for *S. sonnei* (incidence rate ratio 1.24, 95% CI 1.15-1.33), reflecting transmission among MSM. First Nations Australians were disproportionately affected by shigellosis, with the notification rate in this population peaking in 2018 at 92.1 cases per 100,000. The findings of this study provide important insights into the epidemiological characteristics of shigellosis in Australia, and can be used to inform targeted public health prevention and control strategies.

## Introduction

Shigellosis is caused by gram-negative *Shigella* spp. bacteria, which is primarily transmitted through the faecal-oral route by direct or indirect contact with faecal matter (1). There are four species of *Shigella* bacteria, characterised by their O antigen type and biochemical properties: *S. dysenteriae, S. flexneri, S. boydii* and *S. sonnei* (2). Shigellosis outbreaks have been associated with consumption of contaminated food and water; contact with an infected person, particularly in settings with poor hygiene, through sexual contact, or in substandard and crowded living conditions; and recreational contact with contaminated water supplies (3, 4). *Shigella* have a low infectious dose between 10 to 100 organisms, and the clinical presentations of infection can vary from mild watery diarrhoea to severe dysentery (bloody diarrhoea) compounded by systemic complications such as electrolyte imbalance, seizures, fever, nausea and haemolytic uraemic syndrome (5). While shigellosis is often a self-limiting disease, treatment with antibiotics may shorten the duration of the illness and prevent prolonged bacterial shedding, and is recommended for those with severe disease or who are immunocompromised (6, 7). As per Australian guidelines, treatment is also recommended to reduce transmission in key risk groups, including children under the age of six, healthcare workers, and people living or working in residential aged-care facilities, prisons, and other residential facilities (8). Globally, *Shigella* species are a leading cause of diarrhoeal mortality, causing an estimated 200,000 deaths in 2016. (9). The greatest burden of shigellosis is in young children in low- and middle-income countries, where access to good nutrition, clean water, adequate sanitation and healthcare is limited (10). In contrast, the highest rates of shigellosis in high-income countries typically occur in MSM or in travellers returning from overseas (7). Australian national surveillance for shigellosis show a gradual increase in case numbers until 2019, when it more than doubled compared to the 5-year rolling mean (11). Outbreaks have also been reported in First Nations communities in central Australia and among MSM (12, 13).

Stool culture resulting in the isolation of *Shigella* remains the gold-standard method for obtaining a definitive laboratory diagnosis of shigellosis. However, since 2013 there has been an increasing uptake of culture independent diagnostic testing (CIDT) methods such as multiplex polymerase chain reaction (PCR) tests (14). This change in testing practices has implications for public health surveillance of a wide range of enteric pathogens, including *Shigella*. Currently, PCR assays for detecting *Shigella* target the invasion plasmid antigen H (*ipaH)* gene, which is also common to entero-invasive *Escherichia coli* (EIEC) (15). This means that such tests are unable to differentiate between these two organisms, which are closely related and genetically constitute the same species, but typically manifest different symptoms, with EIEC being milder (16). Furthermore, *Shigella* can only be identified to the species and biotype level if an isolate is cultured. Therefore, while PCR testing to detect *Shigella* is more sensitive than traditional bacterial culture methods, the ability to obtain more detailed information, such as antimicrobial susceptibility and epidemiological typing, is compromised. The increasing uptake of CIDT is likely to have implications on the number, accuracy and completeness (with respect to species/biotype data) of surveillance for shigellosis.

In this study, we describe trends in incidence of shigellosis from 2001 to 2019 to understand demographic and geographic factors, including sex, age group, Indigenous status and Australian states and territories. Additionally, we evaluate the impact of CIDT on national surveillance of shigellosis.

## Methods

We analysed shigellosis case notifications made to the National Notifiable Diseases Surveillance System (NNDSS) from 2001 to 2019 by all Australian states and territories. Australia is comprised of eight jurisdictions: New South Wales (NSW), Queensland (QLD), South Australia (SA), Tasmania (TAS), Victoria (VIC) and Western Australia (WA); and two territories: the Australian Capital Territory (ACT) and Northern Territory (NT).

### Data Sources

Shigellosis is a nationally notifiable disease in Australia, and states and territories report all confirmed and probable cases to the Australian Government Department of Health through the NNDSS. The national case definition requires that a confirmed case of shigellosis has *Shigella* species isolated from a clinical specimen; or that *Shigella* is detected by nucleic acid testing in combination with epidemiological evidence (17). The addition of ‘probable’ cases in the surveillance case definition, i.e. detection of *Shigella* by nucleic acid testing only, was implemented from 1 July 2018; prior to this, only confirmed cases were required to be notified. In this study, we did not include ‘probable’ cases.

New South Wales – Australia’s largest state – was the last Australian jurisdiction to make shigellosis reportable, which occurred in 2001. Therefore, we included de-identified data on all confirmed shigellosis case notifications reported to the NNDSS from 2001 to 2019 with the following variables: state; sex; 5 year age-group; month and year of diagnosis; country of acquisition; organism; biotype/subtype; Indigenous status and laboratory diagnosis method. NNDSS data were provided by the Office of Health Protection, Department of Health, on behalf of the Communicable Diseases Network Australia. The month and year of diagnosis was calculated from the diagnosis date: a derived field representing the onset date of the illness or the earliest of the specimen collection date, the date of report, or the date of receipt at the state or territory health authority. Data on laboratory diagnostic method included several categories, including nucleic acid testing, culture, serology, antigen detection, microscopy and unknown. For the purposes of this study, the inputs in this field were consolidated into the following categories depending on whether culture and nucleic acid testing were reported as: PCR; culture; PCR and culture; unknown; and other (any combination of serology, antigen detection, or microscopy that did not also include culture or PCR).

### Analysis

The primary aim of the analysis was to examine the trend in notification rates of shigellosis over time. Cases where sex, age-group and organism were unknown were excluded from the analysis. We calculated rates of illness per 100,000 population using mid-year residential population estimates from the Australian Bureau of Statistics for the years 2001 to 2019.

We used a negative binomial regression model to estimate incidence rate ratios (IRR) by sex, age and state and territory, with significance defined as a p-value of less than 0.05. An interaction term was included in the model to analyse the trend over time for each state and territory, with diagnosis included as a continuous variable. As NSW is the most populous Australian jurisdiction, we applied it as the reference group for the jurisdictional analysis in the model. Due to low (<5) or no cases of *S. dysenteriae* in ACT, NT and TAS, these states were removed from the regression model for *S. dysenteriae*. Stata 15 and Excel 2016 were used for analysis.

### Ethics

We obtained ethical approval for this study from the Australian National University Human Research Ethics Committee [protocol 2018/560] and the ACT Health Human Research Ethics Committee’s Low Risk Sub-Committee [2018/ETH/00158].

## Results

Between 2001 and 2019, Australian states and territories reported 18,363 shigellosis cases to the NNDSS, of which age, sex and organism information were available for >99% (18,327/18,363) cases. Of the cases included in the analysis, 42% (7,649/18,327) were *S. sonnei*, 29% (5,267/18,327) were *S. flexneri*, 1% (214/18,327) were *S. boydii*, and less than 1% (87/18,327) were *S. dysenteriae*. Species information was unknown for 28% (5,110/18,327) of cases, with 79% (4,024/5,110) of these unknown species occurring in the years 2016–2019.

Nationally, crude annual notification rates ranged from 2.2 cases per 100,000 in 2003 and 2011 to 12.4 cases per 100,000 in 2019 (**Error! Reference source not found**.). Rates remained relatively stable between 2001 and 2013, while from 2013 to 2019 there was a greater than five-fold increase in the overall crude notification rate of shigellosis in Australia. Nationally, notification rates increased from 2001 to 2019 with yearly incidence rate ratios of 1.04 (95% CI 1.02-1.07) for *S. boydii*, 1.05 (95% CI 1.04-1.06) for *S. sonnei* and 1.04 (95% CI 1.04-1.06) for *S. flexneri* (Figure 2). The regression model for *S. flexneri* did not fit the data well, which is likely due to outbreaks occurring in the latter years of surveillance in specific states. The IRR for *S. dysenteriae* indicated there was an increase in notification rates between 2001 and 2019, although there was less precision around the interval estimate (IRR 1.01; 95% CI 0.97-1.05).

**Figure 1.**
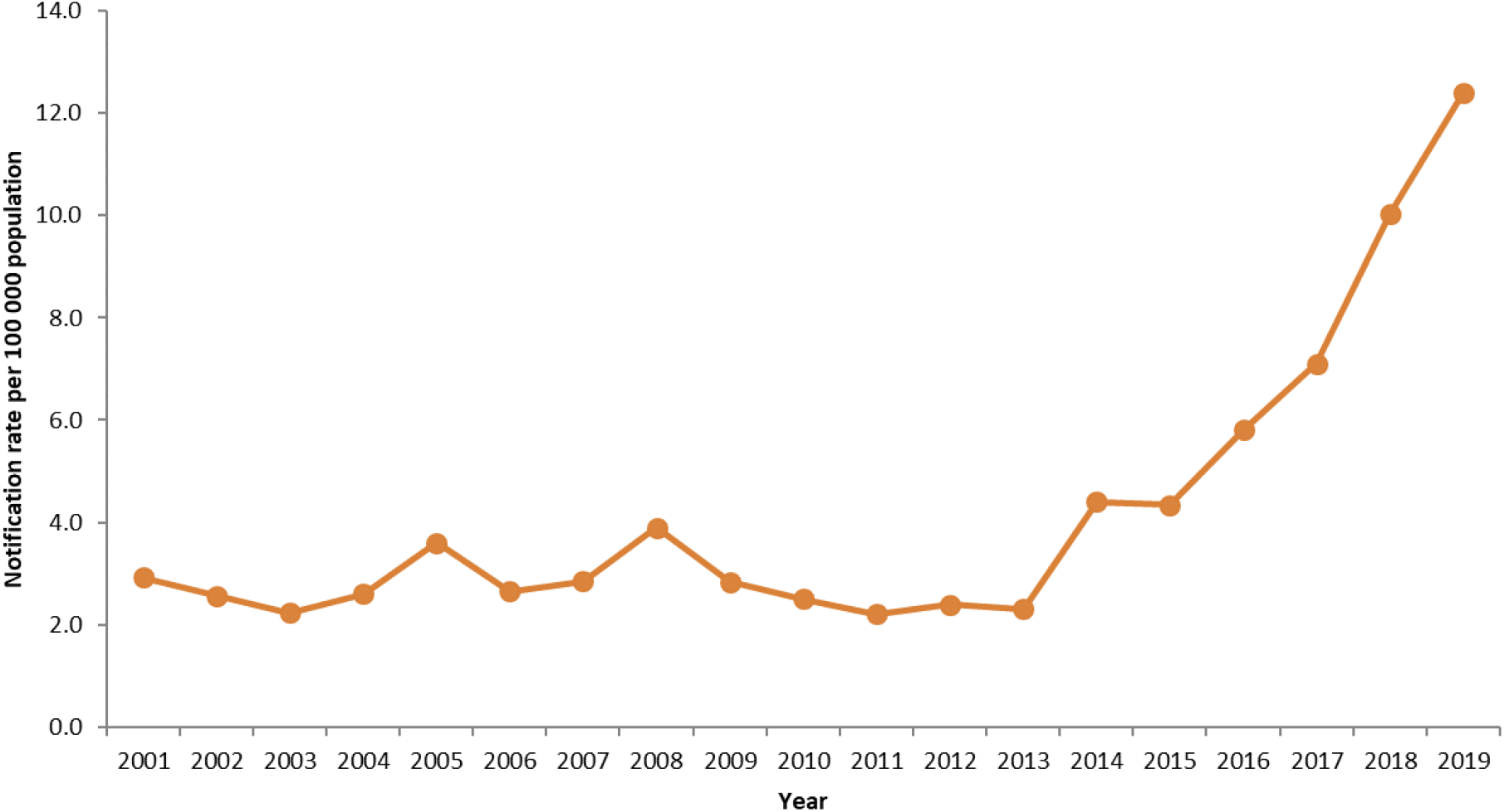
Crude notification rate of shigellosis per 100,000 population, Australia, 2001-2019.

**Figure 2.**
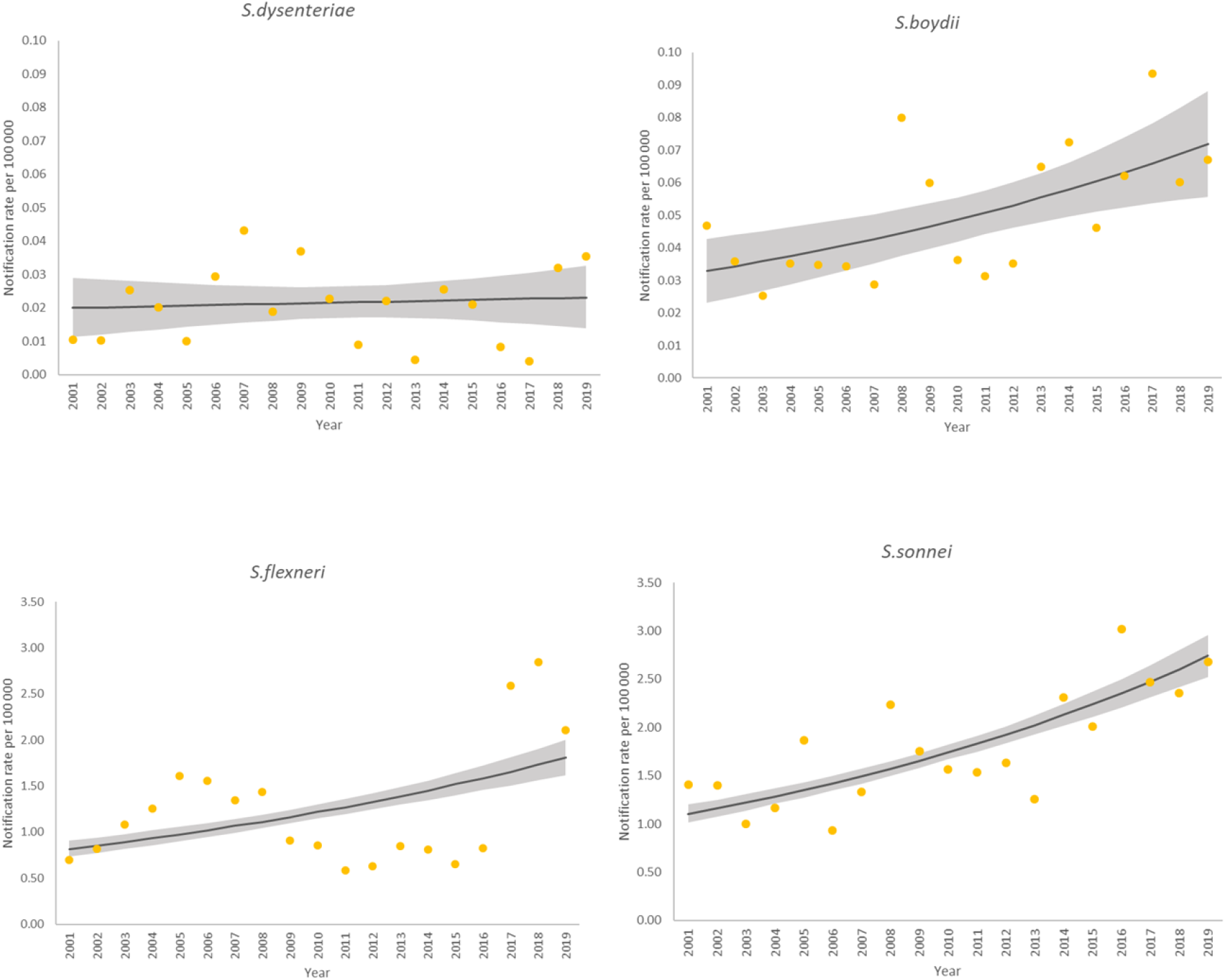
Crude notification rates (dots) and negative binomial regression margins plots (lines with 95% CI) of *S. boydii, S. sonnei, S. flexneri* and *S. dysenteriae*, Australia 2001-2019. *(Note the differing scale of the y-axis for S. boydii and S. dysenteriae, compared to S. sonnei and S. flexneri)*.

Males accounted for 54% (9,844/18,327) of cases. Over the 19-year period, the overall crude notification rate was higher in males than females in the eastern states (ACT, NSW, QLD, and VIC); while the western states and territories experienced higher rates in females (Figure S1). In the state and territory regression model, there was a significantly higher notification rate in males than females for *S. sonnei* (IRR 1.24; 95% CI 1.15-1.33); while there was no significant difference between the sexes for the other species (Table 1).

**Table 1.**
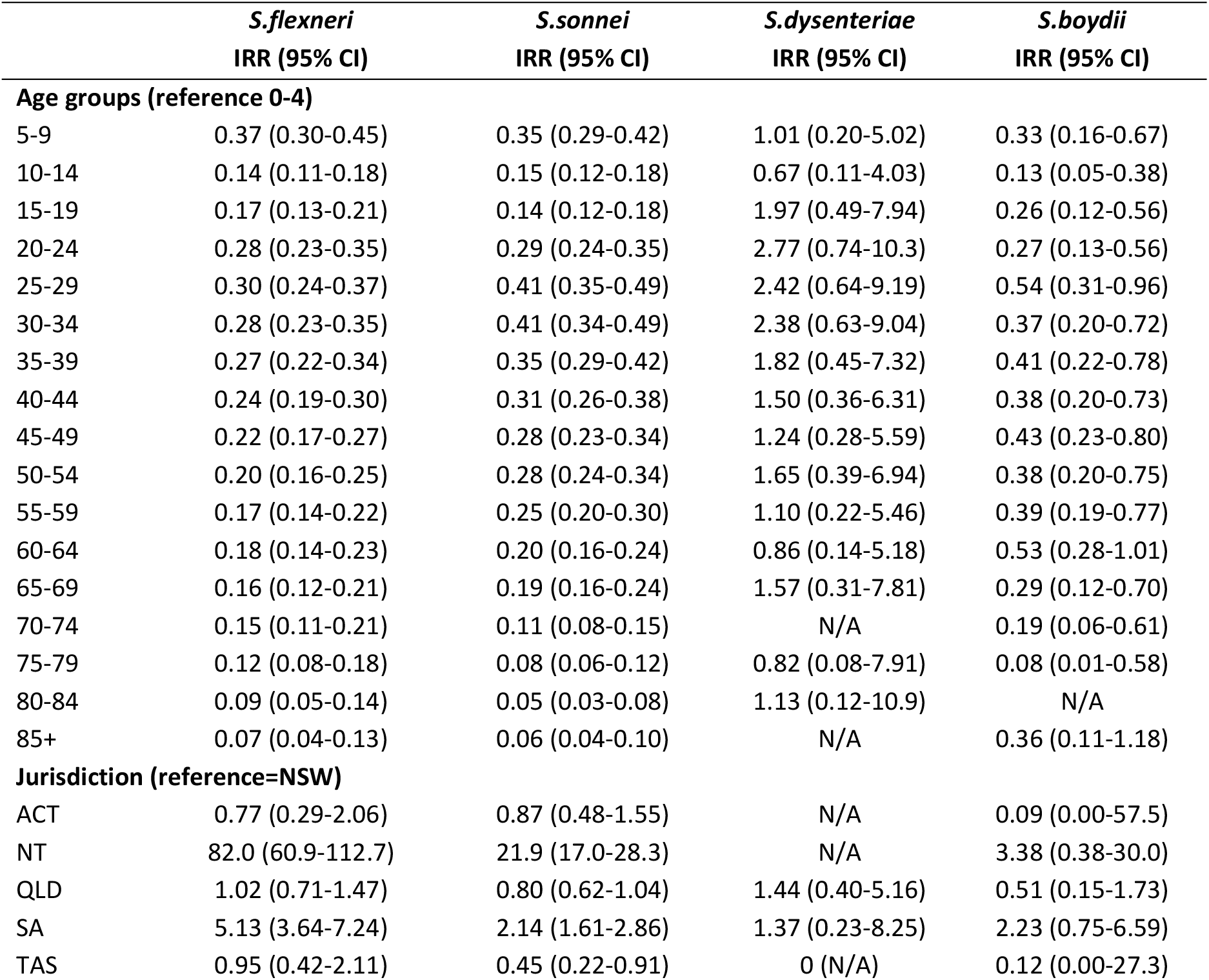
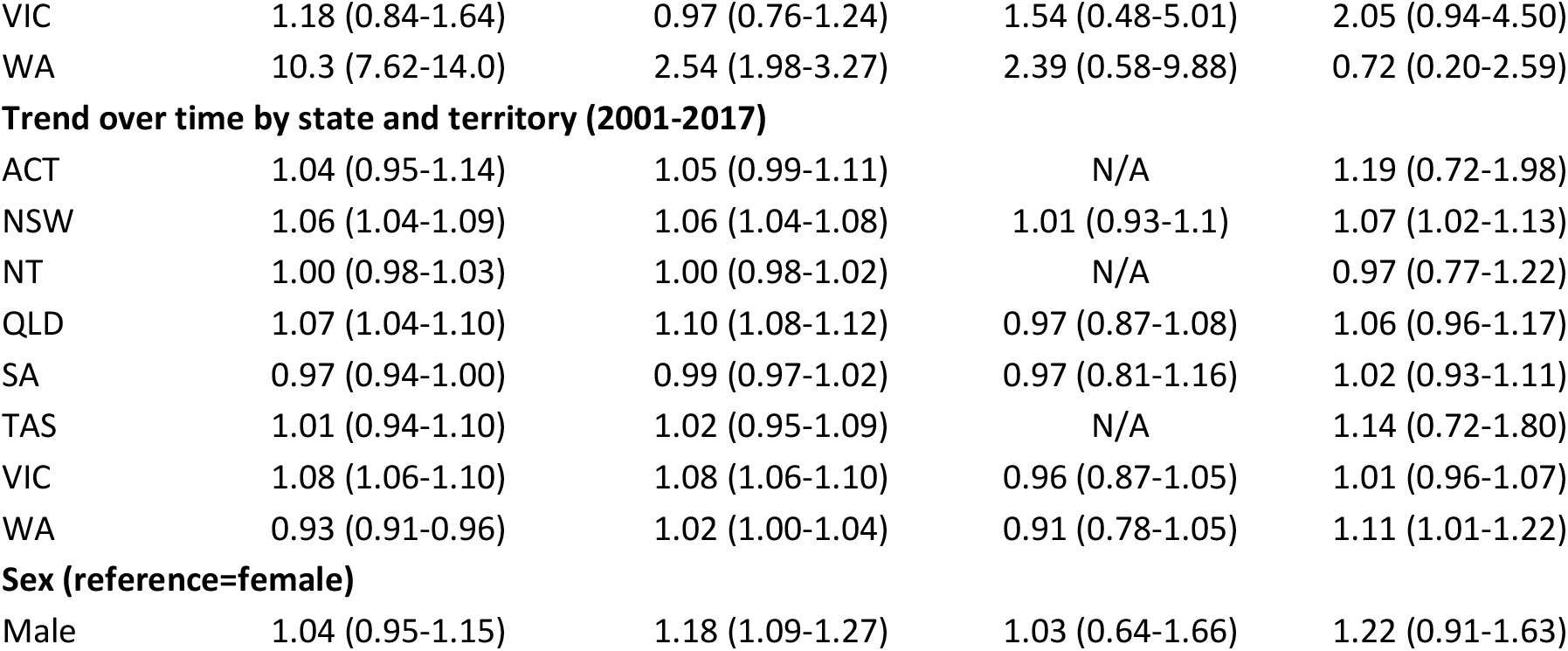
Incidence rate ratios estimated using negative binomial regression of *S. flexneri, S. sonnei, S. dysenteriae* and *S. boydii* by gender, age, state and time, 2001-2019.

|  | *S.flexneri*<br>IRR (95% CI) | *S.sonnei*<br>IRR (95% CI) | *S.dysenteriae*<br>IRR (95% CI) | *S.boydii*<br>IRR (95% CI) |
| --- | --- | --- | --- | --- |
| **Age groups (reference 0-4)** |  |  |  |  |
| 5-9 | 0.37 (0.30-0.45) | 0.35 (0.29-0.42) | 1.01 (0.20-5.02) | 0.33 (0.16-0.67) |
| 10-14 | 0.14 (0.11-0.18) | 0.15 (0.12-0.18) | 0.67 (0.11-4.03) | 0.13 (0.05-0.38) |
| 15-19 | 0.17 (0.13-0.21) | 0.14 (0.12-0.18) | 1.97 (0.49-7.94) | 0.26 (0.12-0.56) |
| 20-24 | 0.28 (0.23-0.35) | 0.29 (0.24-0.35) | 2.77 (0.74-10.3) | 0.27 (0.13-0.56) |
| 25-29 | 0.30 (0.24-0.37) | 0.41 (0.35-0.49) | 2.42 (0.64-9.19) | 0.54 (0.31-0.96) |
| 30-34 | 0.28 (0.23-0.35) | 0.41 (0.34-0.49) | 2.38 (0.63-9.04) | 0.37 (0.20-0.72) |
| 35-39 | 0.27 (0.22-0.34) | 0.35 (0.29-0.42) | 1.82 (0.45-7.32) | 0.41 (0.22-0.78) |
| 40-44 | 0.24 (0.19-0.30) | 0.31 (0.26-0.38) | 1.50 (0.36-6.31) | 0.38 (0.20-0.73) |
| 45-49 | 0.22 (0.17-0.27) | 0.28 (0.23-0.34) | 1.24 (0.28-5.59) | 0.43 (0.23-0.80) |
| 50-54 | 0.20 (0.16-0.25) | 0.28 (0.24-0.34) | 1.65 (0.39-6.94) | 0.38 (0.20-0.75) |
| 55-59 | 0.17 (0.14-0.22) | 0.25 (0.20-0.30) | 1.10 (0.22-5.46) | 0.39 (0.19-0.77) |
| 60-64 | 0.18 (0.14-0.23) | 0.20 (0.16-0.24) | 0.86 (0.14-5.18) | 0.53 (0.28-1.01) |
| 65-69 | 0.16 (0.12-0.21) | 0.19 (0.16-0.24) | 1.57 (0.31-7.81) | 0.29 (0.12-0.70) |
| 70-74 | 0.15 (0.11-0.21) | 0.11 (0.08-0.15) | N/A | 0.19 (0.06-0.61) |
| 75-79 | 0.12 (0.08-0.18) | 0.08 (0.06-0.12) | 0.82 (0.08-7.91) | 0.08 (0.01-0.58) |
| 80-84 | 0.09 (0.05-0.14) | 0.05 (0.03-0.08) | 1.13 (0.12-10.9) | N/A |
| 85+ | 0.07 (0.04-0.13) | 0.06 (0.04-0.10) | N/A | 0.36 (0.11-1.18) |
| **Jurisdiction (reference=NSW)** |  |  |  |  |
| ACT | 0.77 (0.29-2.06) | 0.87 (0.48-1.55) | N/A | 0.09 (0.00-57.5) |
| NT | 82.0 (60.9-112.7) | 21.9 (17.0-28.3) | N/A | 3.38 (0.38-30.0) |
| QLD | 1.02 (0.71-1.47) | 0.80 (0.62-1.04) | 1.44 (0.40-5.16) | 0.51 (0.15-1.73) |
| SA | 5.13 (3.64-7.24) | 2.14 (1.61-2.86) | 1.37 (0.23-8.25) | 2.23 (0.75-6.59) |
| TAS | 0.95 (0.42-2.11) | 0.45 (0.22-0.91) | 0 (N/A) | 0.12 (0.00-27.3) |
| VIC | 1.18 (0.84-1.64) | 0.97 (0.76-1.24) | 1.54 (0.48-5.01) | 2.05 (0.94-4.50) |
| WA | 10.3 (7.62-14.0) | 2.54 (1.98-3.27) | 2.39 (0.58-9.88) | 0.72 (0.20-2.59) |
| Trend over time by state and territory (2001-2017) |  |  |  |  |
| ACT | 1.04 (0.95-1.14) | 1.05 (0.99-1.11) | N/A | 1.19 (0.72-1.98) |
| NSW | 1.06 (1.04-1.09) | 1.06 (1.04-1.08) | 1.01 (0.93-1.1) | 1.07 (1.02-1.13) |
| NT | 1.00 (0.98-1.03) | 1.00 (0.98-1.02) | N/A | 0.97 (0.77-1.22) |
| QLD | 1.07 (1.04-1.10) | 1.10 (1.08-1.12) | 0.97 (0.87-1.08) | 1.06 (0.96-1.17) |
| SA | 0.97 (0.94-1.00) | 0.99 (0.97-1.02) | 0.97 (0.81-1.16) | 1.02 (0.93-1.11) |
| TAS | 1.01 (0.94-1.10) | 1.02 (0.95-1.09) | N/A | 1.14 (0.72-1.80) |
| VIC | 1.08 (1.06-1.10) | 1.08 (1.06-1.10) | 0.96 (0.87-1.05) | 1.01 (0.96-1.07) |
| WA | 0.93 (0.91-0.96) | 1.02 (1.00-1.04) | 0.91 (0.78-1.05) | 1.11 (1.01-1.22) |
| Sex (reference=female) |  |  |  |  |
| Male | 1.04 (0.95-1.15) | 1.18 (1.09-1.27) | 1.03 (0.64-1.66) | 1.22 (0.91-1.63) |

Nationally, notification rates of *S. boydii, S. flexneri* and *S. sonnei* peaked in children aged 0-4 years. The highest rate of cases for *S. dysenteriae* occurred in those aged 20-24 years, however this was not significantly different from the 0-4 year age reference group (IRR 1.40; 95% CI 0.49-3.99). Over the study period, *S. sonnei* was the predominant species in all age groups except in those aged 0-4 years and 80-84 years, where notification rates of *S. flexneri* were slightly higher (Figure 3).

**Figure 3.**
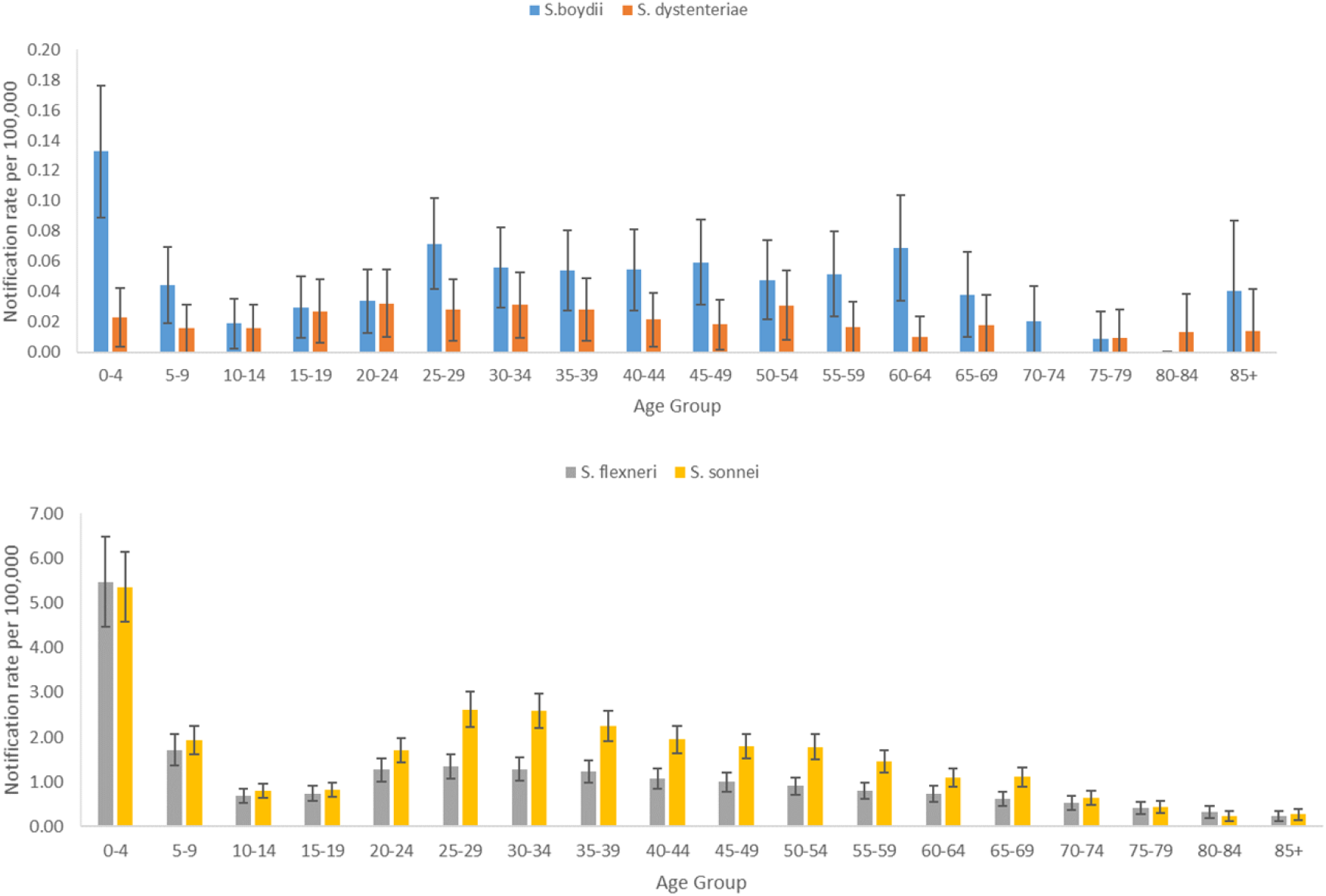
Predicted notification rates of shigellosis per 100,000 with 95% CI, by 5 year age group and species, Australia, 2001-2019. *(Note the differing scale of the y-axis for S. boydii and S. dysenteriae, compared to S. sonnei and S. flexneri)*.

Notification rates varied by jurisdiction, with the NT having considerably higher rates than other states and territories. In 2019, the rate of shigellosis in the NT was 120.3 cases per 100,000 population; nearly 17 times the overall national notification rate (7.1 cases per 100,000 population) (Figure S2). After the NT, in 2019, notification rates were highest in WA (14.8 cases per 100 000 population), followed by NSW (10.6 cases per 100 000 population). At the start of the period, notification rates of *S. flexneri* were significantly higher in the states and territories in the west of Australia, that is NT (IRR 64.87; 95% CI 48.39-86.96), WA (IRR 7.07; 95% CI 5.30-9.44) and SA (IRR 3.61; 95% CI 2.60-5.00), while rates were similar across the eastern states and territories. These western states and territories also had higher rates than the south eastern continental states and territories for *S. sonnei*. The notification rate of this species in TAS was significantly lower than the other jurisdictions (IRR 0.45; 95% CI 0.23-0.88). There was considerable uncertainty in results due to low case numbers making it difficult to conclude there was a difference in the notification rate between states and territories for *S. dysenteriae* and *S. boydii* (Figure 4). The NT, SA and VIC all had higher rates of *S. boydii*, although there was considerable imprecision in the IRR.

**Figure 4.**
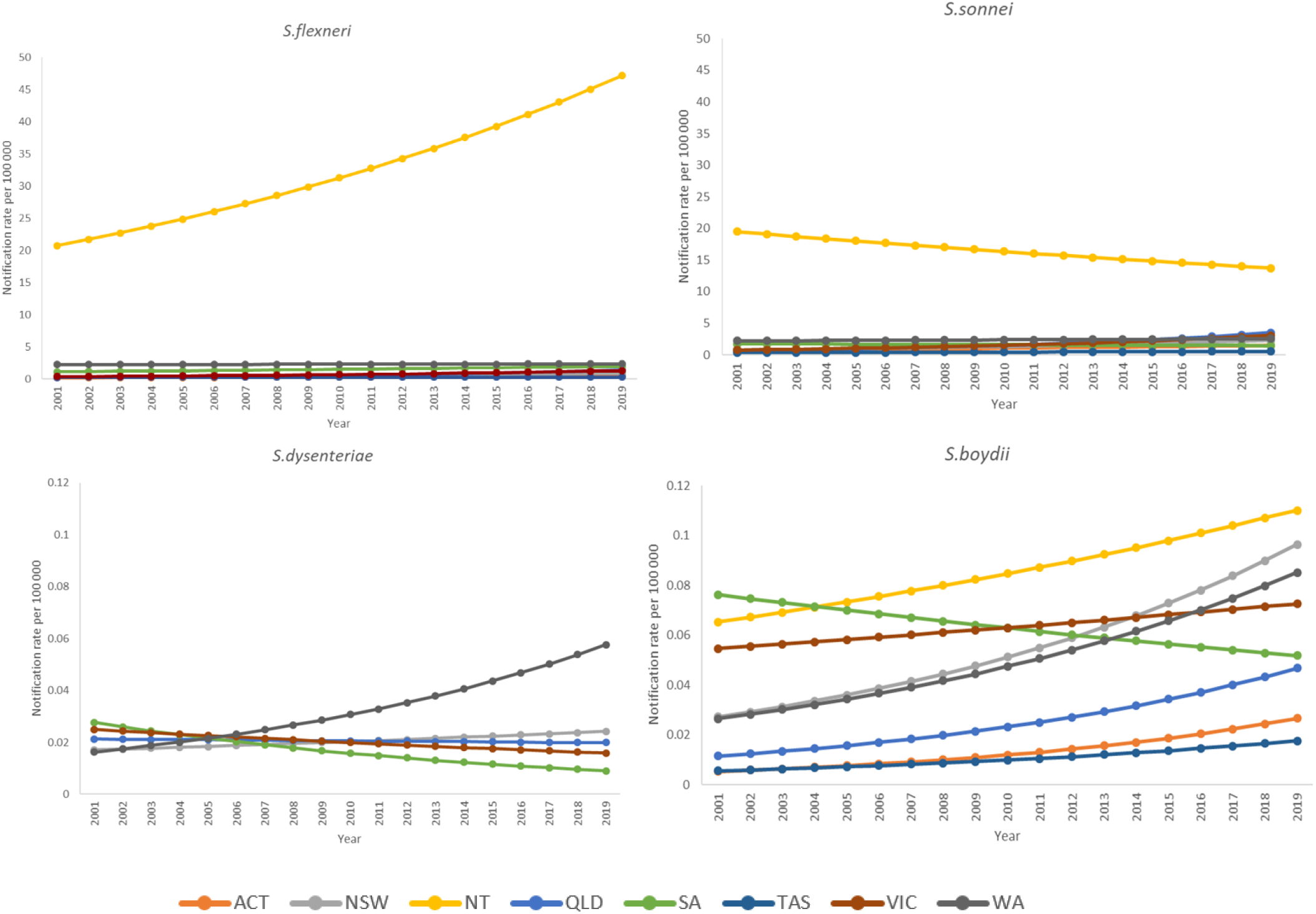
Negative binomial regression margins plot of *S. flexneri, S. sonnei, S. dysenteriae* and *S. boydii* predicted notification rates by state and territory, Australia 2001-2019.

Looking at trends over time, from 2001 to 2019, NSW, NT, QLD, SA and VIC had a significant increase in notification rates for *S. flexneri* (Table 1). Over the same period, notification rates of *S. sonnei* significantly increased in NSW, QLD and VIC, while rates decreased in NT. NSW also experienced a significant increase in *S. boydii*, with an IRR of 1.07 (95% CI 1.03-1.12), along with other jurisdictions although results were imprecise. There was no significant change in the notification rates of *S. dysenteriae* for any state or territory over the 19 year period (Table 1). Regression lines plotted against the crude notification rates for individual state and territories are available in Figure S3.

Indigenous status was available for 82% (15,529/18,327) of cases included in the analysis. There were higher rates of shigellosis among First Nations Australians when compared to the non-Indigenous population (Figure 5). Between 2016 and 2018 there was a greater than 3-fold increase in the notification rate in First Nations Australians, peaking at 92.1 cases per 100,000 in 2018.

**Figure 5.**
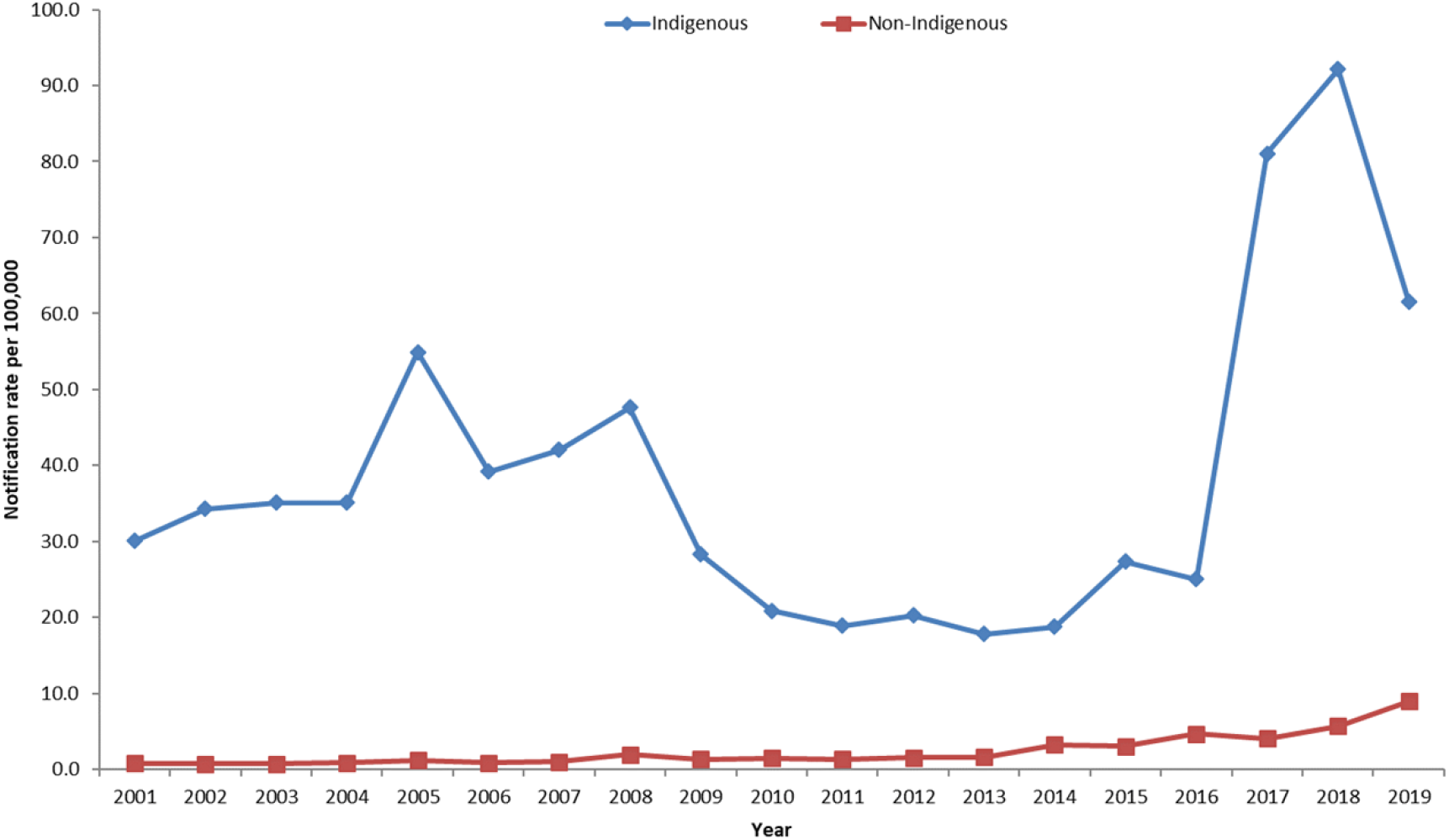
Notification rate of shigellosis per 100,000 population, by Indigenous status, Australia, 2001-2019.

Information on laboratory diagnostic method was available for 87% of cases (16,004/18,327). Between 2001 and 2013, most nationally notified cases of shigellosis were reported to be diagnosed using culture only, peaking at 95% (786/829) of cases in 2008. From 2014 to 2019 there was a decrease in the proportion of cases diagnosed via culture methods, ranging from 54% (554/1,033) in 2014 to 27% (835/3,142) in 2019 due to increasing use of CIDT methods. This led to increased report of untyped *Shigella*, with >63-fold increase in cases of untyped *Shigella* from 2013 (n=30) to 2019 (n=1,902). At a jurisdictional level, all states and territories experienced a dramatic increase in untyped *Shigella* in the past few years (Figure S4).

## Discussion

Notification rates of shigellosis have increased considerably in Australia over the past 19 years (2001-2019), particularly since the introduction of CIDT. The reported rate of all species increased over the time period, except for *S. dysenteriae*. The most common species in Australia was *S. sonnei* (42% all cases), followed by *S. flexneri* (29% of cases), consistent with the reported global burden of shigellosis (10). While *S. flexneri* is traditionally linked to low and middle income countries and *S. sonnei* to high income countries, in recent years the expansion of *S. sonnei* has been documented in many economically transitional regions in Asia, Latin America and the Middle East (18). Globally, data on the epidemiology of *S. boydii* is limited, with this species being predominantly endemic in the Indian Subcontinent (19). The incidence of *S. dysenteriae* in Australia was very low compared to *S. sonnei* and *S. flexneri*, which is consistent with the reported incidence of this species mainly occurring in outbreak settings associated with civil unrest (20). *S. dysenteriae* serotype 1, which is the only *Shigella* species and serotype that expresses the Shiga toxin gene, has high epidemic potential and has been identified as a cause of several large scale outbreaks of dysentery (21-24). This strain is also notorious for developing drug resistance, with epidemic strains typically resistant to multiple antibiotics (25, 26). The epidemiology of *S. dysenteriae* is distinct from the other species of *Shigella*, with epidemics tending to disappear and then reappear years later, with few sporadic cases reported in between outbreaks (27, 28). While large scale outbreaks of *S. dysenteriae* serotype 1 were prevalent in the second half of the twentieth century in Asia, Africa and Central America, there has been a drastic decline in the incidence of *S. dysenteriae* in the last few decades, which remains largely unexplained (5, 20, 29).

The relative importance of *Shigella* species in Australia varied by jurisdiction, with western jurisdictions, particularly the Northern Territory (NT) having higher rates across all years. In Australia, the incidence of shigellosis is higher in First Nations Australians than the non-Indigenous population, which was consistent throughout the period of surveillance. The disparity between the infection rate in First Nations Australians and the non-Indigenous population was particularly pronounced between 2017–2019 due to an outbreak of *S. flexneri* 2b that occurred among First Nations communities in central Australia, predominantly in the NT (12). Antimicrobial resistance has also been reported in endemic strains of *Shigella* affecting First Nations Australians (30). An estimated 18% of First Nations Australians are reported to live in overcrowded households, with the rates of overcrowding highest among those living in remote (26%) and very remote areas (51%) (31). Overcrowding and lack of safe removal and treatment of sewage has shown to be positively associated with gastrointestinal infections in First Nations Australian households (32). A multifaceted and preventative approach is needed to address the disproportional incidence of infectious diseases, including shigellosis, in First Nations communities.

Consistent with global trends, the burden of shigellosis in Australia was highest in children aged less than five years (22, 33). Children in this age group have poor personal hygiene and are yet to have acquired immunity due to a lack of previous exposure (34, 35).

As shigellosis is typically a self-limiting disease, reported cases in NNDSS are likely to underrepresent the true incidence of disease. In general, notification data represent only those cases for which health care was sought, a test conducted, and a diagnosis made, followed by a notification to health authorities. Therefore, the higher incidence of shigellosis in children may also be due to children being overrepresented in the NNDSS dataset due to a higher chance of receiving medical care when unwell, compared to adults. Additionally, the low infectious dose of *Shigella* results in person-to-person transmission, which often occurs between children in childcare settings through close contact (5, 36). Transmission in these settings can also occur due to inadequate handwashing after nappy changing or defecation, or faecal contamination of play areas or nappy changing surfaces (34, 36).

The eastern states of Australia had higher rates of shigellosis in males than females. This is likely to reflect ongoing transmission among MSM in these states with outbreaks occurring over several years in four of the most populous states (QLD, New South Wales, Victoria and South Australia) (37-41).

We identified higher rate of *S. sonnei* in males than females, which is the species responsible for several of the recent outbreaks among MSM in Australian communities. It is vital to control the spread of shigellosis amongst MSM due to the increasing occurrence of drug resistant strains such as multidrug resistant *S. sonnei* biotype G, resistant to all recommended oral antibiotics, which has been reported among MSM in NSW and SA (38, 39).

The shigellosis trends observed in the study are strongly influenced by changes in diagnostic testing methodologies over time. Since 2013, there has been an increase in cases of untyped *Shigella*, coinciding with increased use of culture independent testing methods for shigellosis diagnosis. CIDT are easy to perform, fast, sensitive and reliable, which makes them a valuable diagnostic tool. However, they are generally only able to identify the causative agent of an infection, without providing more granular level information such as pathogen strain, serotype or genotype, which is important to understand disease trends, detect outbreaks and monitor antimicrobial resistance patterns (42). Whole genome sequencing (WGS) allows public health agencies to obtain detailed epidemiological data for surveillance and investigation. WGS-based characterisation of *Shigella* species and serotypes provides higher-resolution and more accurate data than conventional biochemical and serological testing methods. The process to obtain this information is faster as WGS does not require the application of multiple subtyping methodologies (43). Additionally, WGS can differentiate between *Shigella* and EIEC, which have many biochemical properties and virulence genes in common (43, 44). It is important to note however that WGS can only be applied on cultured isolates, which highlights the importance of continuing culture until metagenomics approaches become more available.

Differences between states and territories in the collection of data presents several limitations for this study, particularly in making accurate comparisons between jurisdictions. For instance, the laboratory diagnostic method field in the NNDSS is applied inconsistently between jurisdictions, with some of these applications potentially resulting in an underestimation of cases diagnosed using CIDT and others resulting in an overestimate of cases. Furthermore, prior to the implementation of the new case definition in July 2018, testing and notification practices varied by jurisdiction. The old shigellosis case definition defined laboratory definitive evidence as ‘*isolation or detection of Shigella species*’. Based on this, most states interpreted a PCR positive result as not a detection and waited for a positive result from reflex culture before making a notification. However, some states sent all these case notifications to the NNDSS marked as ‘probable’, which were recorded by the NNDSS system as ‘confirmed’ cases. Therefore, the observed increase in shigellosis cases since 2013 may be in part attributable to changes in testing (increased PCR uptake) and notification practices due to inconsistent interpretations of the case definition by states and territories. The differing testing practices between jurisdictions is evident when looking at the prevalence of untyped *Shigella*. For instance, VIC, NT and Tasmania all had a pronounced increase in the proportion of untyped *Shigella* from 2014 onwards, which is attributable to the uptake of PCR testing in these jurisdictions.

Returning travellers are a high risk group for shigellosis (40). To examine this trend on a national level, data on the place of acquisition of shigellosis cases was collected for this study. However the lack of completeness of this field limited its reliability for analysis and meaningful interpretation.

## Conclusion

Since 2001, there has been an increasing incidence of shigellosis in Australia, with CIDT methods and persistent outbreaks in specific population groups responsible for much of the increase. This study has contributed to improving the understanding of the characteristics of shigellosis activity, including identifying high risk groups such as young children, MSM and First Nations Australians. The study also observed a considerable increase in untyped *Shigella* in recent years which coincided with an increased use of CIDT. This highlights the implications of culture independent testing on shigellosis surveillance, with CIDT leading to a reduction in the availability of species level information. Enhancements in surveillance and laboratory practices, such as through the application of the revised surveillance case definition and use of WGS, will provide further insights into the epidemiological features of shigellosis in Australia, thereby enabling the implementation of targeted and efficient prevention and control interventions.

## Data Availability

The data underlying the results presented in the study are from the National Notifiable Diseases Surveillance System. As the data was obtained from a third party, we are unable to upload the minimal data set used in this study. Australian shigellosis notification data, as was used in this study, can be requested from the Communicable Diseases Network Australia. Data requests can be sent to.

